# Unbiased characterization of atrial fibrillation phenotypic architecture provides insight to genetic liability and clinically relevant outcomes

**DOI:** 10.1101/2024.02.13.24302788

**Authors:** Giovanni Davogustto, Shilin Zhao, Yajing Li, Eric Farber-Eger, Brandon D. Lowery, Lauren Lee Shaffer, Jonathan D. Mosley, M. Benjamin Shoemaker, Yaomin Xu, Dan M. Roden, Quinn S. Wells

**Affiliations:** Department of Medicine, Division of Cardiovascular Medicine, Vanderbilt University Medical Center, Nashville TN; Department of Biostatistics, Vanderbilt University Medical Center, Nashville TN; Department of Biomedical Informatics, Vanderbilt University Medical Center, Nashville TN

## Abstract

**Background:** Atrial Fibrillation (AF) is a common and clinically heterogeneous arrythmia. Machine learning (ML) algorithms can define data-driven disease subtypes in an unbiased fashion, but whether the AF subgroups defined in this way align with underlying mechanisms, such as high polygenic liability to AF or inflammation, and associate with clinical outcomes is unclear.

**Methods:** We identified individuals with AF in a large biobank linked to electronic health records (EHR) and genome-wide genotyping. The phenotypic architecture in the AF cohort was defined using principal component analysis of 35 expertly curated and uncorrelated clinical features. We applied an unsupervised co-clustering machine learning algorithm to the 35 features to identify distinct phenotypic AF clusters. The clinical inflammatory status of the clusters was defined using measured biomarkers (CRP, ESR, WBC, Neutrophil %, Platelet count, RDW) within 6 months of first AF mention in the EHR. Polygenic risk scores (PRS) for AF and cytokine levels were used to assess genetic liability of clusters to AF and inflammation, respectively. Clinical outcomes were collected from EHR up to the last medical contact.

**Results:** The analysis included 23,271 subjects with AF, of which 6,023 had available genome-wide genotyping. The machine learning algorithm identified 3 phenotypic clusters that were distinguished by increasing prevalence of comorbidities, particularly renal dysfunction, and coronary artery disease. Polygenic liability to AF across clusters was highest in the low comorbidity cluster. Clinically measured inflammatory biomarkers were highest in the high comorbid cluster, while there was no difference between groups in genetically predicted levels of inflammatory biomarkers. Subgroup assignment was associated with multiple clinical outcomes including mortality, stroke, bleeding, and use of cardiac implantable electronic devices after AF diagnosis.

**Conclusion:** Patient subgroups identified by unsupervised clustering were distinguished by comorbidity burden and associated with risk of clinically important outcomes. Polygenic liability to AF across clusters was greatest in the low comorbidity subgroup. Clinical inflammation, as reflected by measured biomarkers, was lowest in the subgroup with lowest comorbidities. However, there were no differences in genetically predicted levels of inflammatory biomarkers, suggesting associations between AF and inflammation is driven by acquired comorbidities rather than genetic predisposition.

## INTRODUCTION

Atrial fibrillation (AF) is the most common arrhythmia encountered in clinical practice,^1^ and is associated with increased mortality,^2^ stroke,^3^ heart failure (HF),^3^ myocardial infarction,^4,5^ dementia,^6^ and decreased quality of life.^7^ AF is a complex, dynamic, and heterogeneous condition, and there is considerable variation in the age at onset, arrhythmia burden, presence of risk factors, severity of symptoms, and incidence of complications among affected individuals.^8–18^ Heterogeneity is likewise evident in response to AF treatments, including antiarrhythmic drugs and catheter-based ablation.^19,20^ This interindividual variation suggests complex gene-environment interactions and the existence of multiple AF subtypes with differing mechanistic underpinnings.

Two sources of risk that may contribute to AF heterogeneity, and by extension, AF subgroups, are genetic susceptibility to AF and inflammation, which itself may be due to acquired or genetic factors. Indeed, polygenic liability conferred by common variants is associated with risk of AF and multiple clinical outcomes.^21–26^ Additionally, multiple lines of evidence support a significant role for inflammation in the pathophysiology of AF. For example, elevated levels of inflammatory biomarkers^27^ and polymorphisms in genes encoding inflammatory cytokines^28–31^ are associated with AF risk; and inflammatory infiltrates are observed in atria of AF patients.^32^ Moreover, many AF risk factors (e.g., HF, coronary artery disease [CAD], hypertension [HTN], obesity) and biologic mechanisms (e.g., fibrosis, thrombogenesis) are themselves associated with increased inflammation,^25,26,33^ and AF itself can promote inflammation and perpetuate atrial remodeling and arrhythmia.^34,35^ However, the contributions of genetic susceptibility to AF and inflammation, both acquired and genetically driven, to AF phenotypic subgroups has not been reported.

The emergence of advanced machine learning (ML) methods has facilitated data-driven subtyping of complex, heterogeneous cardiovascular diseases such as HTN,^36^ HF,^37^ and pulmonary arterial hypertension.^38^ ML-derived phenotypic AF subgroups have been identified in post hoc analyses in observational registries.^39,40^ These studies, however, did not explore the mechanistic or genetic basis of subgroups. Therefore, there is no consensus regarding the classification of AF subtypes based on their underlying mechanisms, particularly with respect to patients in clinical care.^41,42^

We hypothesized that contemporary ML methods applied to a large clinical AF population will identify distinct subtypes with different polygenic liability of AF, inflammatory state, and clinical outcomes.

## MATERIALS AND METHODS

The study protocol was approved by the Vanderbilt University Medical Center (VUMC) Institutional Review Board (VUMC IRB# 181403).

### Data source

Clinical data were derived from the Synthetic Derivative (SD), a de-identified version of the Vanderbilt University Medical Center’s Electronic Health Record (EHR) intended to support research.^43–45^ For the subset of participants with genetic information, genotype data were obtained from BioVU, the Vanderbilt biobank linked to the SD that contains DNA samples and genetic data derived from clinical blood samples that would otherwise be discarded.^46,47^

### Study population

Subjects with AF were identified using a random-forest classifier applied to routinely collected clinical features. A list of features, including International Classification of Diseases codes (ICD codes), Current Procedural Terminology (CPT) codes, text strings, and medications were used in the classifier (**Table S1**). Every instance of each feature was identified and timestamped across the EHR for each subject. The instance/date pairs were used to generate 3 scoring metrics for each feature: strength (number of distinct days with a mention), persistence (number of days between first and last mention of a feature), and durability (ratio between feature persistence and total record length). The date of the first qualifying feature (e.g., code or text mention) for atrial fibrillation was used as date of AF first recognition for each subject. The classifier was trained and validated locally and demonstrated superior performance compared to conventional phenotyping approaches such as ICD codes. Analyses were limited to subjects receiving ongoing care at our institution, defined as ≥3 outpatient visits to primary care or cardiology over 5 years. Clustering was based on feature values at the time of AF first recognition in the EHR. Subjects with missing predictors or outcome data were excluded. When available, genome-wide genotyping data for subjects was obtained from BioVU. Genetic analyses were restricted to subjects of European and African ancestries due to the small number (n=48) of subjects from other ancestries with genotype data. A summary of inclusion and exclusion criteria is presented in **Figure S1**.

### Predictor features

Clustering used 35 clinical features selected as relevant for AF by two cardiologists (GD, QSW), and included demographics, body mass index (BMI), elements of the CHA2DS2-VASc score, sleep apnea, presence of cardiac implantable electronic devices, left ventricular systolic dysfunction, thyroid disease, and valvular heart disease. Features were extracted from the de-identified EHR using a variety of techniques including queries of demographics, anthropometric measures, diagnosis and procedure codes, and natural language processing of clinical notes and procedure reports. A description of features and definitions is presented in Supplemental File 1. Only features prior to or on the day of AF first recognition were utilized for clustering. An exception was made for BMI due to sparsity of weight/height measures, where the closest value to AF first recognition - before or after - was used. As described below, an initial pairwise analysis confirmed that the 35 selected features were not themselves highly correlated.

### Outcomes

Clinical outcomes included mortality, bleeding-related hospitalization, stroke, pacemaker implantation, implantable cardioverter defibrillator (ICD) implantation, cardioversion, and AF ablation. Inflammatory burden in clusters was assessed using clinical inflammatory biomarker levels ascertained within 6 months of first AF mention. Polygenic risk scores, derived from published GWAS, were used to assess genetic liability to AF and imputed inflammatory cytokine levels (further details below). Outcomes were required to first appear in the medical record at least one day after the first mention of AF. A full description of outcomes and definitions is presented in Supplemental File 2.

### Genetic data, AF polygenic risk score calculation, and imputed cytokine calculation

Genotyping data for BioVU subjects were acquired using the Illumina Multi-Ethnic Genotyping Array (MEGA) platform. QC was performed using PLINK v1.90α6.17, and included duplicate removal, sex-check, and removal of one of each pair of related individuals (pi-hat>0.2). Data were then pre-processed using the HRC-1000G-check tool v4.2.11 and imputed using the Michigan Imputation server and the 10/2014 release of the 1,000 Genomes cosmopolitan reference haplotypes.

Genetic predisposition to AF within clusters was assessed using a previously published polygenic risk score (PRS) that comprises 1,168 SNPs (https://www.pgscatalog.org/score/PGS000035/).^48^ In our dataset, 8 SNPs did not pass QC and hence the PRS value was computed by summing the product of allele weighting and allele dosage across 1,160 available SNPs. The raw scores were then normalized to the sample mean and standard deviation. The SNPs and associated weights included in the AF PRS are presented in Supplemental File 3.

Genetic predisposition for inflammation between clusters was assessed using genetically predicted cytokine levels, as previously described by Mosley et al.^49^ Briefly, using published GWAS summary statistics of measured cytokine levels, we calculated a PRS value (“imputed cytokine level”) of BioVU subjects for each cytokine of interest with at least one SNP meeting genome-wide significance (GWS) in published summary statistics.^49^ Again, PRS were computed by summing the product of allele weighting and allele dosage across the SNPs passing GWS.

Imputed cytokine levels of 41 cytokines that met inclusion criteria were created. To validate that imputed cytokine levels were capturing a clinically relevant predisposition to disease, we conducted phenome-wide association studies (PheWAS) for each cytokine PRS in Vanderbilt’s de-identified EHR. Of 41 imputed cytokines, 13 had at least one significant association (FDR <0.1) with clinical phenotype and were used in downstream analyses. A summary of imputed cytokines selection is presented in Figure S2, and the results of validation PheWAS of all the included cytokines are available for download in the supplemental material.

### Clustering

Due to requirements of the clustering algorithm, continuous variables such as age and BMI were converted into categories. Age was categorized into <50, 50-70, and > 70 years at first AF mention in EHR. BMI was categorized into <18 Kg/m^2^, 18-24.99 Kg/m^2^, 25-29.99 Kg/m^2^, 30-39.99 Kg/m^2^, and >=40 Kg/m^2^. Transformed data were analyzed using the BlockCluster package, employing the coclusterBinary function specifically designed for binary data, and therefore all feature elements were coded in the form of dummy variables. The clustering methodology requires the number of clusters to be prespecified, which in this case was set to 3 clusters based on the distribution of features. The clustering algorithm involves estimating a latent mixture block model, and simultaneously conducts block clustering in both subject and features sets. The number of clusters were prespecified to 3 based on the number of features used.

### Other statistical considerations and analyses

Statistical analyses were performed with the R statistical program (v4.3.1), and primarily using the “Blockcluster” (4.5.3), “Hmisc” (v5.1.1), and “rms” (v6.7.1) packages. Unless otherwise specified, continuous variables are presented as median (interquartile range), and categorical variables as counts (percentage). Differences in measured inflammatory biomarkers among clusters were assessed using one way ANOVA. The associations of cluster assignment with normalized AF PRS and imputed cytokine levels from genetic data were determined using an ordinal regression model. The models tested the association between PRS as continuous variables and cluster assignment, and included age, sex, and 4 principal components of ancestry as covariates. For all these sets of tests, false discovery rates (FDRs) were calculated from the nominal *p-values* using the R function p.adjust using the Benjamini & Hochberg option. FDRs less than 0.1 were considered statistically significant. A Cox proportional hazards regression model was used to test the association of cluster assignment and time-to-event mortality, adjusting for age of first AF mention and CHA_2_DS_2_-VASc score. The functions *adjustedsurv* and *adjusted_surv_quantile* of the package adjustedCurves (v0.10.1) were used to create the adjusted survival curves and adjusted median survival estimates. For other outcomes, unadjusted cox proportional hazards regression models were used to test the association of cluster assignment and time-to-event outcomes.

## RESULTS

### Cohort characteristics

We identified 23,271 subjects with AF meeting inclusion criteria. The cohort median age was 68 years (IQR: 59-76), 42.6% female, and 90.4% self-reported White. The most prevalent comorbidity was hypertension (62.3%), followed by renal dysfunction (36.8%), coronary artery disease (CAD) (35.8%), and fluid-electrolyte disorders (25.9%). Heart failure was also common (18.4%), and 12.1% of patients had history of left ventricular systolic dysfunction (Table 1).

**Table 1:** Baseline characteristics of the included participants at time of first AF mention in EHR. Age, BMI, and CHA_2_DS_2_-VASc score are presented as median (IQR). All other variables are categorical or dichotomous presented as n (%). Abbreviations: ICD: Implantable Cardioverter Defibrillator, EHR: Electronic Health Record.

| Characteristic | All participants (n=23,271) |
| --- | --- |
| Age, years | 68 (59-76) |
| Female Sex | 9,908 (42.6) |
| Self-reported Race |  |
| White | 21,032 (90.4) |
| Black | 1,782 (7.7) |
| Asian | 177 (0.8) |
| Native American | 36 (0.2) |
| Islander | 31 (0.1) |
| Unknown | 213 (0.9) |
| Ethnicity |  |
| Non-Hispanic | 22,873 (98.3) |
| Hispanic | 156 (0.7) |
| Unknown | 242 (1.0) |
| BMI, Kg/m<sup>2</sup> | 29 (25.2-33.8) |
| CHA<sub>2</sub>DS<sub>2</sub>-VASc score | 3 (2-4) |
| Diabetes mellitus | 3,621 (15.6) |
| Hypertension | 14,500 (62.3) |
| Thyroid medication use | 4,264 (18.3) |
| Obstructive sleep apnea | 2,562 (11.0) |
| Rheumatologic diseases | 2,971 (12.8) |
| Chronic Obstructive Pulmonary Disease | 1,811 (7.8) |
| Prior stroke | 1,093 (4.7) |
| Congestive heart failure | 4,281 (18.4) |
| Left ventricular systolic dysfunction | 2,805 (12.1) |
| Coronary artery disease | 8,332 (35.8) |
| Valvular heart disease | 2,200 (9.5) |
| Prior pacemaker | 1,748 (7.5) |
| Prior ICD | 1,411 (6.1) |
| Renal dysfunction | 8,559 (36.8) |
| Peripheral arterial disease | 1,272 (5.5) |
| Pulmonary circulatory disorders | 2,312 (9.9) |
| Paralysis | 418 (1.8) |
| Other neurological disorders | 1,024 (4.4) |
| Liver disease | 1,519 (6.5) |
| Solid tumor | 2,052 (8.8) |
| Coagulopathy | 2,433 (10.5) |
| Weight loss | 351 (1.5) |
| Fluid electrolyte disorders | 6,024 (25.9) |
| Blood loss anemia | 494 (2.1) |
| Deficiency anemia | 1,392 (6.0) |
| Alcohol abuse | 169 (0.7) |
| Drug abuse | 453 (1.9) |
| Psychoses | 76 (0.3) |
| Depression | 2,585 (11.1) |

The median follow-up after AF first mention was 5.4 (IQR:2.8-8.6) years. Mortality during follow-up was 14%, with a median time to death of 4.5 years (IQR 1.6-7.8) after first AF mention. Approximately half of subjects (55.2%) received antiarrhythmic drug therapy, usually initiated shortly after diagnosis (median 14.6 days, IQR 0.0-401.5). Cardioversion was performed in approximately one fifth of the subjects (19.9%) and AF ablation in 8.1%. Device placement was common, with pacemaker implantation in 13.7% of subjects and 8.5% undergoing ICD implantation. Stroke occurred in 12.3% of subjects while bleeding events were rare (Table 2).

**Table 2:** Clinically relevant incident outcomes of the included participants after first AF mention in EHR.

| **Outcome** | **All participants (n=23,271)** |
| --- | --- |
| Follow up, years | 5.4 (2.8-8.6) |
| Dead, n (%) | 3,259 (14.0) |
| Time to death, years | 4.5 (1.6-7.8) |
| Stroke after AF diagnosis, n (%) | 2,861 (12.3) |
| Time to first stroke, years | 1.6 (0.1-5.1) |
| Bleeding related hospitalization, n (%) | 784 (3.4) |
| Bleeding source, n (%) |  |
| Intracranial | 294 (1.3) |
| Gastrointestinal | 276 (1.2) |
| Genitourinary | 69 (0.3) |
| Other | 145 (0.6) |
| Time to initial bleeding related hospitalization, years | 2.6 (0.6-5.4) |
| Traumatic bleeding hospitalization, n (%) | 333 (1.4) |
| Traumatic bleeding source, n (%) |  |
| Gastrointestinal | 295 (1.3) |
| Intracranial | 20 (0.1) |
| Genitourinary | 2 (0) |
| Other | 16 (0.1) |
| Time to initial traumatic bleeding hospitalization, years | 2.3 (0.6-5.7) |
| AF ablation, n (%) | 1,884 (8.1) |
| Time to initial AF ablation, years | 2.4 (0.6-5.5) |
| Cardioversion, n (%) | 4630 (19.9) |
| Time to initial cardioversion, years | 0.3 (0.0-3.0) |
| Antiarrhythmic drug therapy, n (%) | 12,847 (55.2) |
| Time to initial antiarrhythmic, years | 0.04 (0.0-1.1) |
| Pacemaker after AF diagnosis, n (%) | 3,184 (13.7) |
| Time to pacemaker implant, years | 1.48 (0.2-4.2) |
| ICD after AF diagnosis, n (%) | 1,972 (8.5) |
| Time to ICD implant, years | 1.3 (0.2-4.4) |

Abbreviations: ICD: Implantable Cardioverter Defibrillator, EHR: Electronic Health Record.

### Feature exploration

To gain additional insight into the phenotypic architecture of our cohort prior to clustering, we conducted a series of exploratory analyses. We first assessed pairwise correlations between features to identify patterns among variables, detect pairs of highly correlated features that could affect clustering, and inform *a priori* estimates of the expected number of groups derived by clustering. The selected features were not highly correlated (all r<0.5), and all were retained for clustering analyses. Based on initial visual evaluation, we estimated that clustering of would result in approximately 3 distinct phenotypic subgroups based on correlation clustering (Figure 1A). We then conducted data dimensionality reduction using principal component analysis (PCA) and evaluation of variable contributions to components using a loading plot (Figure 1B). This revealed that comorbidity burden, particularly left ventricular systolic dysfunction, fluid-electrolyte disorders, coronary artery disease, and renal dysfunction, were the main contributors to principal component (PC) 1, while age group was the main contributor for PC2. Analysis of PC associations with mortality revealed that PC1 values were associated with increased risk of death (PC1 interquartile OR 1.99, 95% CI: 1.91-2.07).

**Fig. 1:**
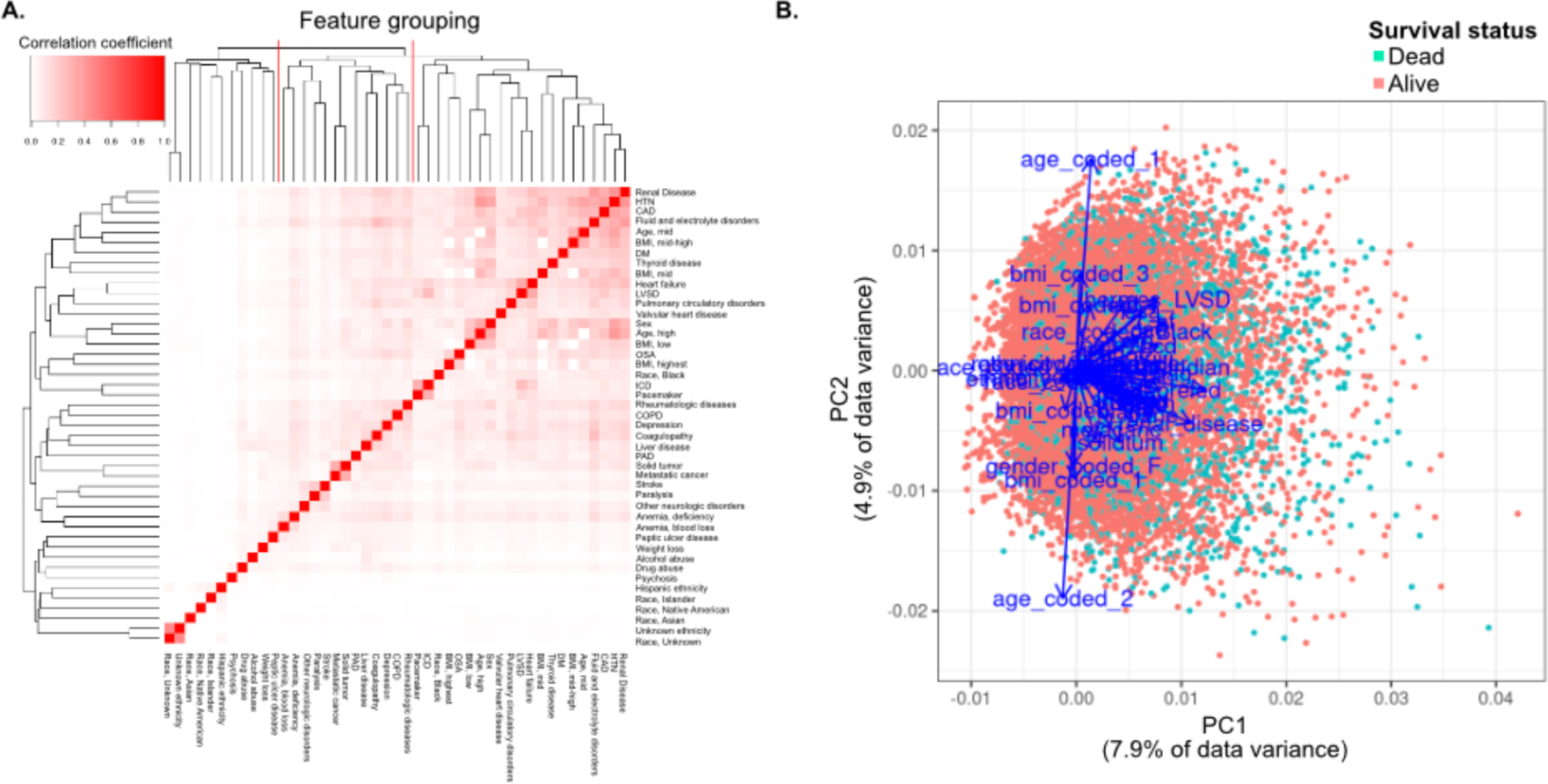
Overall exploration, and correlation of clinical variables used as predictors in clustering algorithm. A) Observed correlation between clinical predictors used for clustering. There were no highly correlated features (r<0.5). Correlation clustering of features suggest approximately 3 groups (separated by vertical red lines in top horizontal axis) B) Principal component analysis of predictors reveal PC1 is mainly determined by comorbidity burden, while PC2 is mostly determined by age at time of AF diagnosis and BMI. There is an association between positive PC1 and death during follow up (p<0.001). Abbreviations: AF: Atrial fibrillation, CAD: coronary artery disease, BMI: Body mass index, COPD: Chronic obstructive pulmonary disease, PAD: Peripheral arterial disease, LVSD: Left ventricular systolic dysfunction, ICD: Implantable cardioverter defibrillator, OSA: Obstructive sleep apnea, DM: Diabetes mellitus, HTN: Hypertension, PC: Principal component.

### Unbiased clustering of features at AF diagnosis

The co-clustering algorithm *blockcluster* groups subjects and features simultaneously to identify blocks of subjects with similar characteristics. Based on results above, 3 clusters of subjects were prespecified, but also identified 3 distinct groups of predictors determining cluster assignment. Clusters were distinguished by increasing burden of comorbidities (Figure 2A), particularly renal dysfunction, fluid-electrolyte disorders, and CAD, as shown in Figure 2B. Based on comorbidity burden, we defined groups as “Low”, “Mid”, and “High” comorbidity clusters. Figure S3 shows that other comorbidities demonstrated similar patterns of distribution across clusters, but the magnitude was less pronounced than renal dysfunction, coronary artery disease, and fluid-electrolyte disorders (Figure S3).

**Fig. 2:**
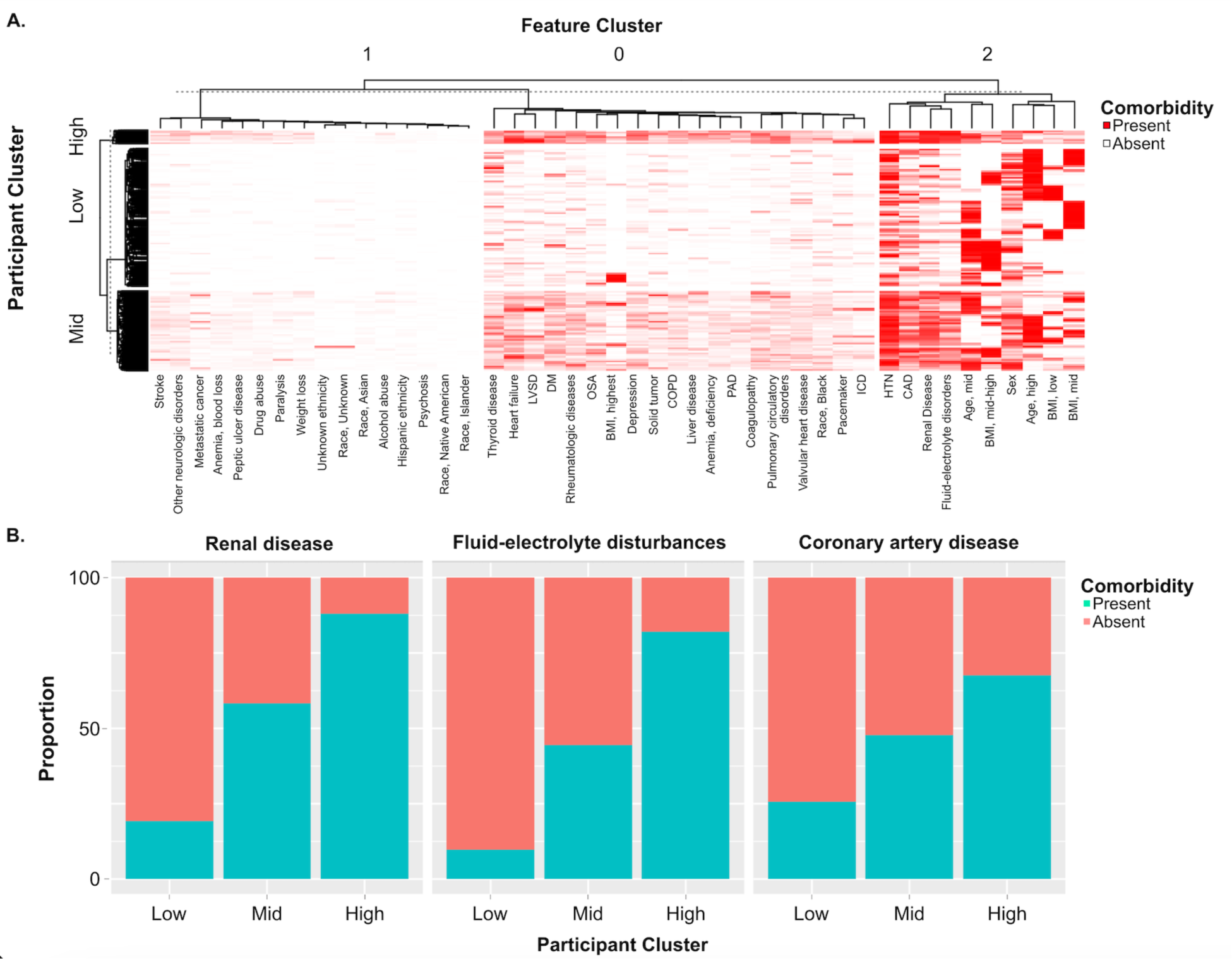
Clustering assignment and feature prevalence at AF diagnosis time by cluster. A) Heatmap of cluster assignment by participant and feature. Each row represents a participant, and each column a feature. White denotes absence of feature at AF first mention time while red presence of feature. Renal dysfunction, fluid-electrolyte disorders, and CAD are the most striking features which determine participant cluster assignment. B) Comorbidity burden for renal dysfunction, fluid-electrolyte disorders, and CAD distribution across clusters. Abbreviations: AF: Atrial fibrillation, VHD: Valvular heart disease, CAD: coronary artery disease, BMI: Body mass index, COPD: Chronic obstructive pulmonary disease, PAD: Peripheral arterial disease, LVSD: Left ventricular systolic dysfunction, ICD: Implantable cardioverter defibrillator, OSA: Obstructive sleep apnea, DM: Diabetes mellitus, HTN: Hypertension, Rheum: Rheumatologic diseases

### Association of clusters with clinical outcomes

Unadjusted survival differed significantly among clusters (Median survival 23.2 years [95% CI 22-greater than 25] for Low cluster, 16.7 years [95% CI 15.1-18.5] for Mid cluster, and 9.5 years [95% CI 8.3-10.9] for High cluster), with a dose-response effect observed between increasing cluster comorbidity burden and worse survival (Figure 3A). A similar relationship was seen with mortality when the 3 key variables determining cluster assignment (renal dysfunction, CAD, fluid-electrolyte disorders) were considered (Figure 3B, Median survival >25 years when all these variables were absent, 20.2 years [95% CI 19.1-22.3] with one variable present, and 13.6 years [95% CI 12.5-15.1] with two or three variables present). Furthermore, when adjusting for age and CHA2DS2-VASc score at the time of first AF mention, cluster assignment remained significantly associated with survival (adjusted median survival 22.02 years [95% CI 21.34-23.17] for Low cluster, 13.98 years [95% CI 13.51-14.53] for Mid cluster, and 7.96 years [95% CI 7.39-8.64] for High cluster, Figure 3C).

**Fig. 3:**
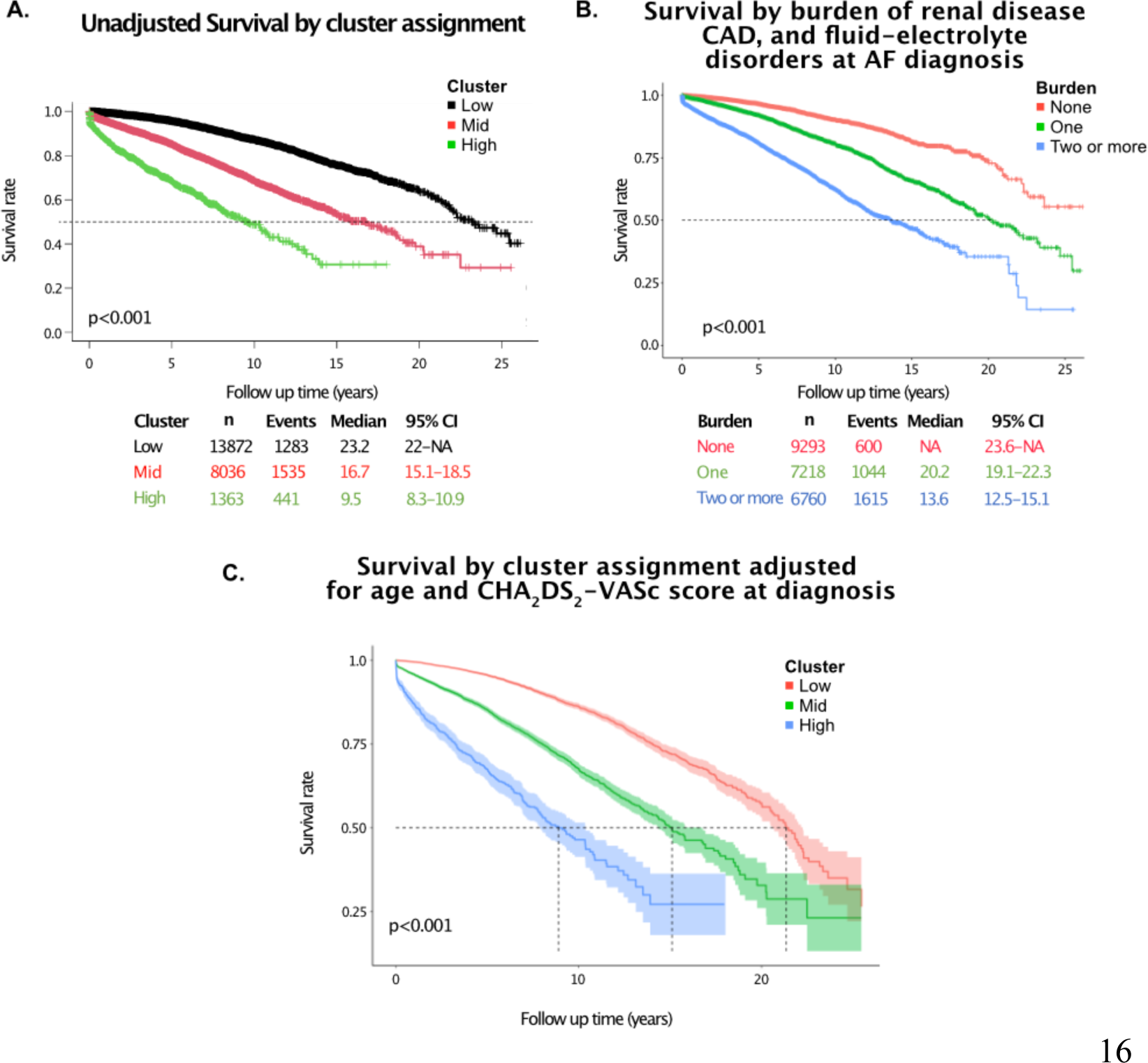
Survival by cluster assignment and by burden of renal dysfunction, coronary artery disease, and fluid-electrolyte disorders. A) Kaplan-Meier curves demonstrating observed survival by cluster assignment. B) Kaplan-Meier curves demonstrating observed survival by burden of renal dysfunction, coronary artery disease (CAD), and fluid-electrolyte disorders. C) Predicted survival by cluster assignment adjusted for age at CHA2DS2-VASc score at time of first AF mention. The solid line represents point-estimate and the shaded area the 95% point-wise confidence interval. Abbreviations: AF: Atrial fibrillation, CAD: coronary artery disease.

Like survival, there was a significant difference in unadjusted event-rates for stroke, bleeding, pacemaker placement, ICD placement, ablation, and cardioversion. Again, clusters with greater burden of comorbidities had more events observed for these outcomes (Figure 4, figure S4). In general, patients in the “High” cluster were more likely to have episodes of stroke or bleeding and ICD implantation, while patients in the “Low” cluster were more likely to have ablations and cardioversions. Patients in the “Mid” cluster were most likely to have pacemaker implantation. Antiarrhythmic drug therapy was comparable across clusters (Figure S4).

**Fig. 4:**
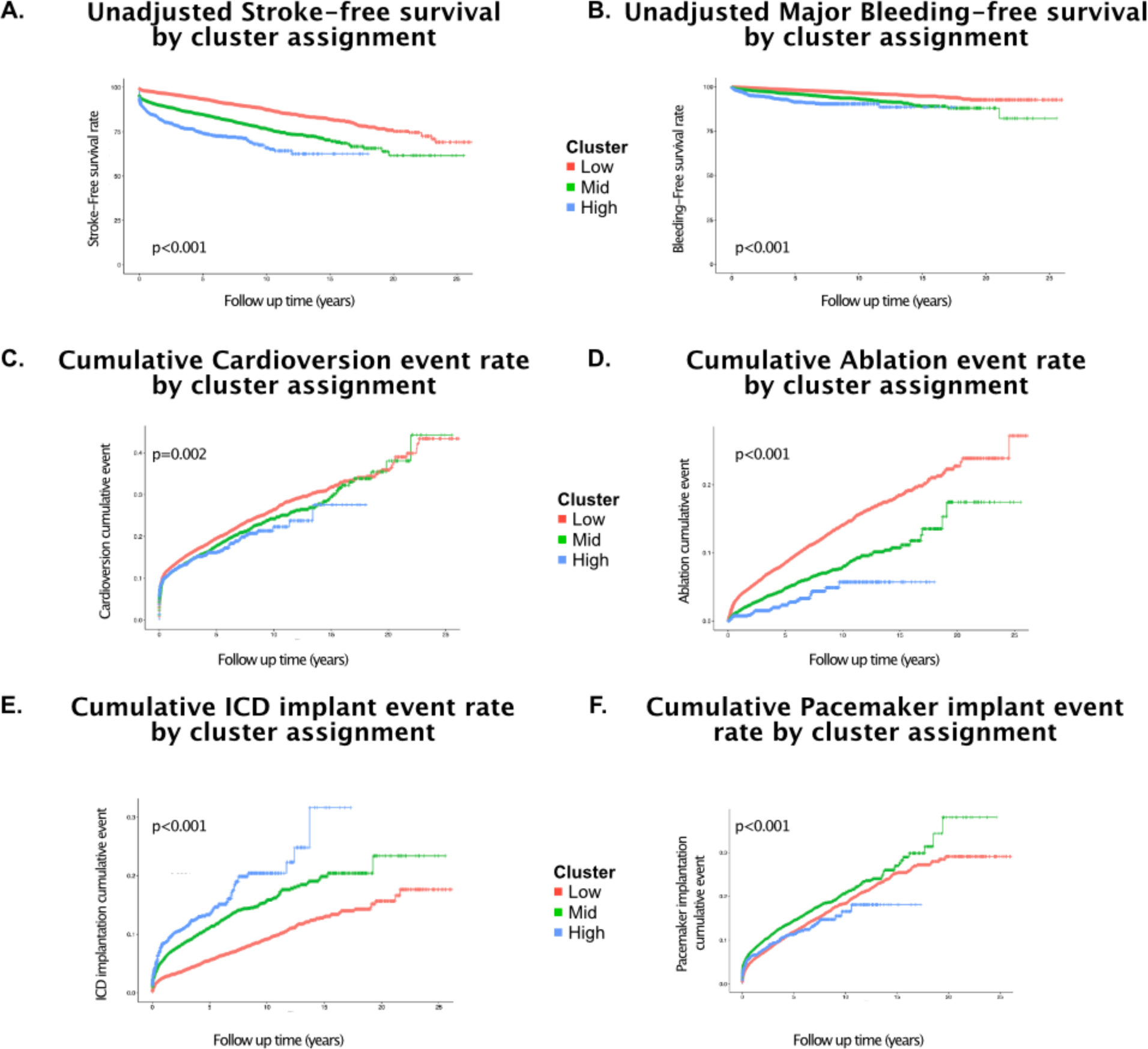
Unadjusted clinical outcomes during follow up by cluster assignment. A) Kaplan-Meier curves demonstrating observed stroke-free survival by cluster assignment. B) Kaplan-Meier curves demonstrating observed major bleeding-free survival by cluster assignment. C) Observed cumulative cardioversion event rate by cluster assignment. D) Observed cumulative atrial fibrillation ablation event rate by cluster assignment. E) Observed cumulative ICD implantation event rate by cluster assignment. C) Observed cumulative pacemaker implantation event rate by cluster assignment. Abbreviations: AF: Atrial fibrillation, ICD: Implantable cardioverter-defibrillator.

### Association of clusters with AF polygenic risk, measured inflammatory biomarkers, and imputed cytokine levels

There was a significant difference among clusters with respect to genetic liability to AF assessed by normalized AF PRS after adjustment for age, sex, and first 4 principal components of ancestry, with the highest genetic susceptibility to AF seen in the “Low” comorbidity cluster and lowest genetic susceptibility to AF in the “High” comorbidity clusters (Figure 5).

**Fig. 5:**
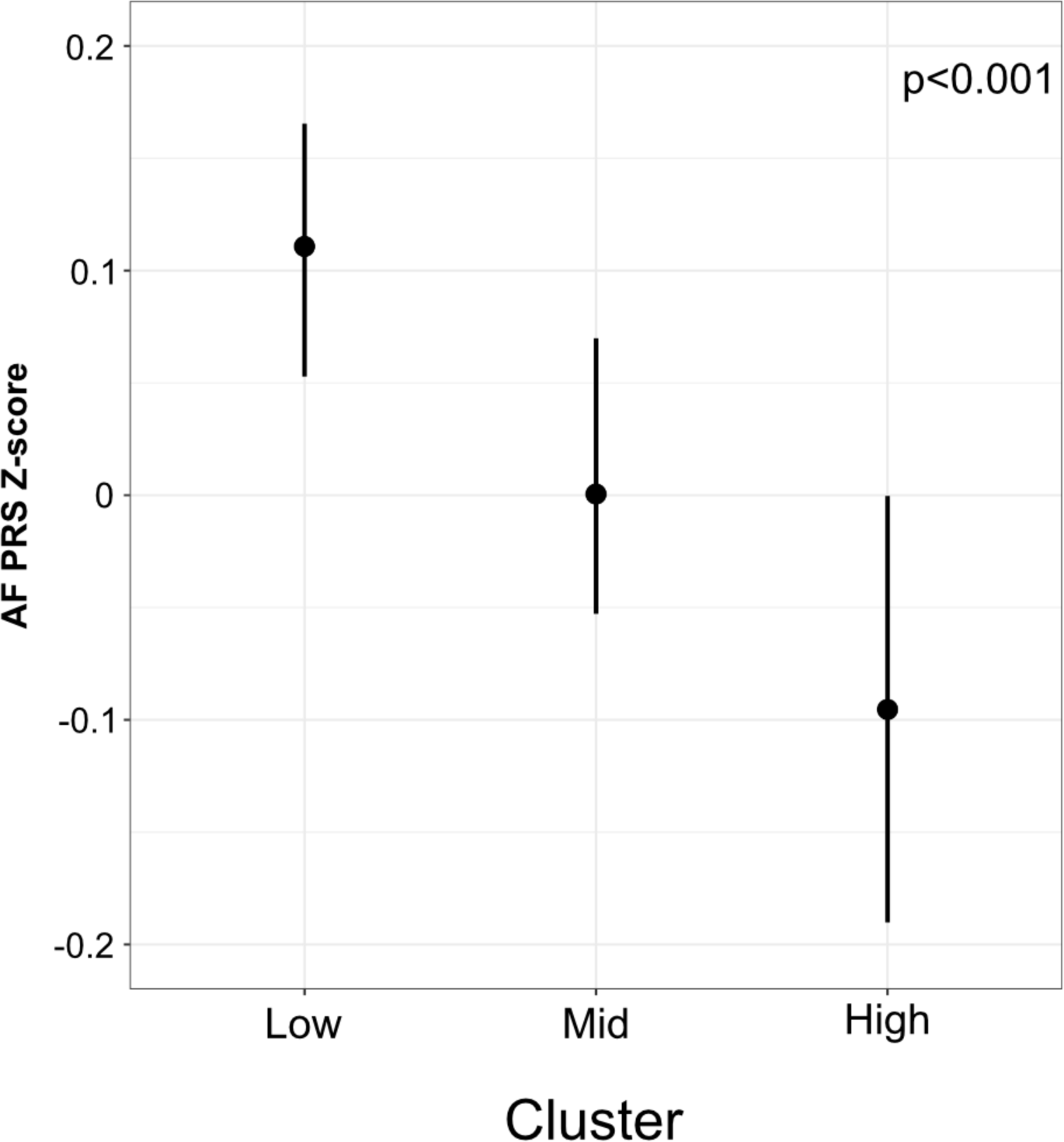
Normalized atrial fibrillation polygenic risk score by cluster adjusted for age, sex, and first 4 principal components of ancestry. Normalized (Z-score) of atrial fibrillation polygenic risk score (AF PRS) by cluster group adjusted for first 4 principal components of ancestry, age at first AF mention, and sex. Data are presented as median predicted values after adjustment (dot) and pointwise 95% confidence interval (bars). Abbreviations: AF: Atrial fibrillation, PRS: Polygenic risk score.

We compared clusters with respect to clinically ascertained levels of C-reactive protein, erythrocyte sedimentation rate, and white blood cell count measured within 6 months of AF first mention. There was a significant association between AF cluster and measured inflammatory biomarkers (Table 3): all biomarkers (p<0.001 for each) had lowest values in the “Low” comorbidity cluster and highest values in the “High” comorbidity cluster, with intermediate values in “Mid” cluster.

**Table 3:** Inflammatory biomarkers levels by cluster.

| Biomarker | Cluster |  |  |  | p-value |
| --- | --- | --- | --- | --- | --- |
|  | Overall | Low | Mid | High |  |
| Erythrocyte sedimentation rate | 28 (12-53)<br>n=2,206 | 19 (8.3-42)<br>n=855 | 32 (14.4-56)<br>n=1,052 | 42 (21-69.5)<br>n=299 | <0.001 |
| C-reactive protein (mg/L) | 21 (4.7-85.6)<br>n=2,174 | 11.5 (2.8-57.4)<br>n=639 | 26.2 (5.5-93.72)<br>n=1,164 | 34.2 (9.5-102)<br>n=371 | <0.001 |
| Platelet count | 212 (168-261.5)<br>n=15,771 | 213 (173-261)<br>n=7,851 | 212 (165-262.3)<br>n=6,619 | 206 (153-261)<br>n=1,301 | <0.001 |
| White blood cell count | 7.9 (6.3-9.8)<br>n=16,438 | 7.6 (6.1-9.5)<br>n=8,339 | 8.2 (6.5-10.2)<br>n=6,790 | 8.2 (6.5-10.2)<br>n=1,309 | <0.001 |
| Neutrophil percentage | 66.5 (58.3-74.5)<br>n=11,533 | 64.7 (56.8-72.7)<br>n=5,146 | 67.7 (59.5-74.5)<br>n=5,210 | 69.4 (61.5-76.7)<br>n=1,177 | <0.001 |
| Red cell distribution width | 14.2 (13.3-15.4)<br>n=16,335 | 13.8 (13.1-14.7)<br>n=8,268 | 14.6 (13.6-15.9)<br>n=6,759 | 15.5 (14.3-17)<br>n=1,308 | <0.001 |

Data are presented in median (IQR), and all comparisons made with one-way ANOVA test.

The PheWAS of genetically predicted circulating cytokines identified 13 markers (IFN-GR1, IL-10RB, IL-17F, IL-17RD, IL-1RL2, IL-1RN, IL-23R, IL-34, IL-37, IL6R, IL-6ST, TNFRSF-11B, TNFRSF-6B) to take forward in this analysis. After accounting for multiple comparison testing, there were no significant differences among clusters with respect to genetically imputed levels for these inflammatory cytokines and cytokine-associated circulating proteins. The normalized imputed cytokine levels values overall and by cluster are presented in Supplemental File 4.

## DISCUSSION

We conducted an analysis of 23,271 subjects with atrial fibrillation from a large tertiary medical center that identified 3 phenotypic disease clusters associated with mortality, stroke, and bleeding. Cluster membership was largely driven by comorbid diseases, in particular the presence of renal dysfunction, CAD, and fluid-electrolyte disturbances. Greater comorbidity burden across groups was associated with higher levels of clinically measured inflammatory biomarkers and inversely associated with a polygenic predisposition for AF. There was no detectable difference among groups with respect to genetic predisposition to inflammation as ascertained by imputed cytokine levels.

Many of our findings are in agreement with prior studies reporting data-driven subgroups of AF, and thereby expands and strengthens the existing evidence base.^39,40^ For example, two earlier studies (n= 9,749 and 2,458 subjects, respectively), both using observational cohorts and hierarchical clustering, identified 3-4 clusters, including clusters comprising subjects with young age or low comorbidity burden and a cluster with CAD as driving feature. Additionally, both studies reported associations between cluster assignment and major adverse cardiovascular events. Like these studies, we observed a similar number of clusters, with CAD and other comorbidities (renal dysfunction and fluid-electrolyte disturbances) as key drivers of cluster assignment and an association of cluster assignment with mortality, stroke, bleeding, cardioversion, ablation, and cardiac implantable electronic device (CIED) implant. The present study adds to the evidence base of data-driven analyses of AF phenotypic architecture and supports the importance of comorbidities as determinates of interindividual variability in AF outcomes.

Our work also extends previous work in important ways. One notable distinction from prior studies is the size of the study population, which comprised over 23,000 subjects from a large medical center with dense phenotype data and more than 5 years of follow-up after AF first mention in the EHR. This population is more than twice the size of earlier reports and was sufficiently large to enable more detailed analyses of AF phenotypic architecture including evaluation of a broader range of comorbidities, inflammatory markers, and polygenic risk. For example, we identified renal dysfunction as an important determinate of cluster assignment, a finding that has not been previously reported. This observation warrants replication in other studies but suggests a previously underappreciated contribution of renal dysfunction to AF phenotypic heterogeneity, especially in clinical populations.

The current report also provides new insights into potential mechanisms underlying AF subgroups, including clinically acquired and genetically determined inflammation as well as genetic susceptibility to AF itself. We observed higher levels of clinically measured inflammatory biomarkers in clusters with more comorbidities. However, there were no differences with respect to genetically predicted levels of inflammatory cytokines. This suggests that observed epidemiologic associations between inflammation and AF are driven by acquired comorbidities and/or environmental factors rather than a genetic predisposition to increased inflammation. These observations could explain, at least in part, the lack of universal efficacy of anti-inflammatory agents as AF therapies^50–53^, and further support the importance of cardiovascular risk factor optimization in the management of AF.

We also observed greater polygenic risk for AF in groups with lower comorbidity burden, with genetic risk highest in the lowest burden of CAD and other comorbidities. Clinically, this cluster has many similarities with a group of patients previously referred to as having “lone AF”. Collectively, in the context of prior literature, our findings suggest that observed phenotypic variability in AF, in part, is distributed along a gradient with differing contributions of genetic risk and acquired comorbidity. This framework for understanding AF heterogeneity, incorporating both environmental and genetic factors, aligns with clinical experience and has been previously articulated in a theoretical context.^54^ However, to our knowledge, our analysis is the first to empirically demonstrate the complementary roles of comorbidity and genetic liability as major drivers of phenotypic variability using hypothesis-free methods.

### Limitations

Even though we took steps to improve quality and completeness of EHR-derived data, including restricting the analysis to subjects receiving longitudinal care in our health system, misclassification and under-ascertainment of clinical phenotypes and outcomes, notably mortality which may be under-estimated in the EHR, are possible due to misdiagnosis or underdiagnosis. Additionally, measured biomarkers were not prospectively collected and therefore are susceptible to ascertainment bias. Only a portion of patients included had genetic data available which limited the power of the imputed cytokine analyses. It is also possible that imputed cytokines did not capture sufficient genetic variability of cytokine levels; we attempted to minimize this issue by only using imputed cytokines validated by phenome-wide scanning in the analyses.

## Conclusions

Patient subgroups identified by unsupervised clustering were associated with differing risk of mortality, stroke, bleeding, and ICD implant. These groups were distinguished by comorbidity burden and polygenic liability to AF, which were inversely correlated. Clinical inflammation, as reflected by higher measured biomarker levels, was associated with increasing comorbidity burden, but there were no differences in genetically predicted levels of inflammatory biomarkers, consistent with the idea that inflammatory burden is driven by acquired comorbidities.

## Data Availability

Data used in this study are available upon request to the corresponding author and after approval from institutional review.

## Acknowledgements

Clinical data were obtained using Vanderbilt’s Synthetic Derivative. The Synthetic Derivative resource is supported by CTSA award No. UL1TR000445 from the National Center for Advancing Translational Sciences. Genomic data were obtained using Vanderbilt’s de-identified biobank BioVU. Vanderbilt University Medical Center’s BioVU projects are supported by numerous sources: institutional funding, private agencies, and federal grants. These include NIH funded Shared Instrumentation Grant S10OD017985, S10RR025141, and S10OD025092; CTSA grants UL1TR002243, UL1TR000445, and UL1RR024975.The contents of this publication are solely the responsibility of the authors and do not necessarily represent official views of the National Center for Advancing Translational Sciences or the National Institutes of Health.

## Sources of Funding

This project was supported by Vanderbilt’s participation in the American Heart Association’s Atrial Fibrillation Strategically Focused Research Network (18SFRN34110369 and 18SFRN34230089). This project also received support from NIH grants R01 HL149826 (DMR), R01 HL142856 (JDM).

## Disclosures

None.

## Notes

### Competing Interest Statement

The authors have declared no competing interest.

### Funding Statement

This project was supported by Vanderbilt participation in the American Heart Associations Atrial Fibrillation Strategically Focused Research Network (18SFRN34110369 and 18SFRN34230089). This project also received support from NIH grants R01 HL149826 (DMR) and R01 HL142856 (JDM).

### Author Declarations

Vanderbilt University Medical Center (VUMC) Institutional Review Board (IRB#181403)

