## Supplement for "Unbiased characterization of atrial fibrillation phenotypic architecture provides insight to genetic liability and clinically relevant outcomes"

**Table of Contents**

**Table S1:** Features used in the AF random forest classifier.

**Figure S1:** Atrial fibrillation cohort selection and genetic subsample.

**Figure S2:** Imputed cytokine selection and validation process.

**Figure S3:** Feature prevalence at AF diagnosis time by cluster.

**Figure S4:** Clinical outcomes during follow up by cluster.

**Table S1: Features used in the AF random forest classifier**

| Feature group | Definition |
|---|---|
| ICD9 codes | 427.31 |
| ICD10 codes | I48.00*, I48.91*, I48.1*, and I48.2* |
| CPT codes | 93656, 93657, 93620, 93621, 92960, 92961 |
| Free-text terms | "atrial fibrillation", "pulmonary vein isolation", "cardioversion", "afib", "atrial fib", "atrial-fib", "a fib", "DCCV", "AF" |
| Medications | Quinidine, Procainamide, Disopyramide, Mexiletine, Propafenone, Flecainide, Dronedarone, Amiodarone, Sotalol, Ibutilide, Dofetilide, Verapamil, Diltiazem, Mexitil, Rythmol, Tambocor, Multaq, Pacerone, Betapace, Corvert, Tikosyn, Nexterone |

**Figure S1: Atrial fibrillation cohort selection and genetic subsample**

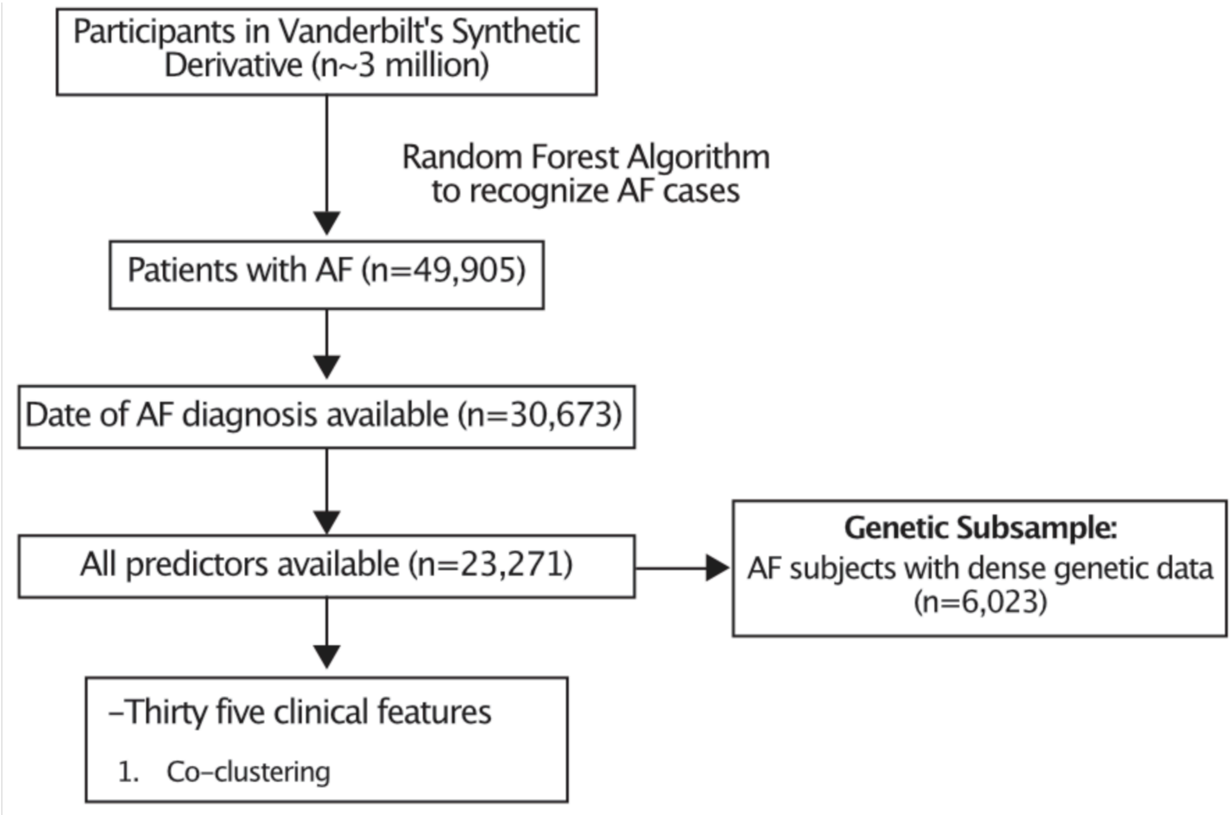


Abbreviations: AF: atrial fibrillation.

**Fig. S2: Imputed cytokine selection and validation process**

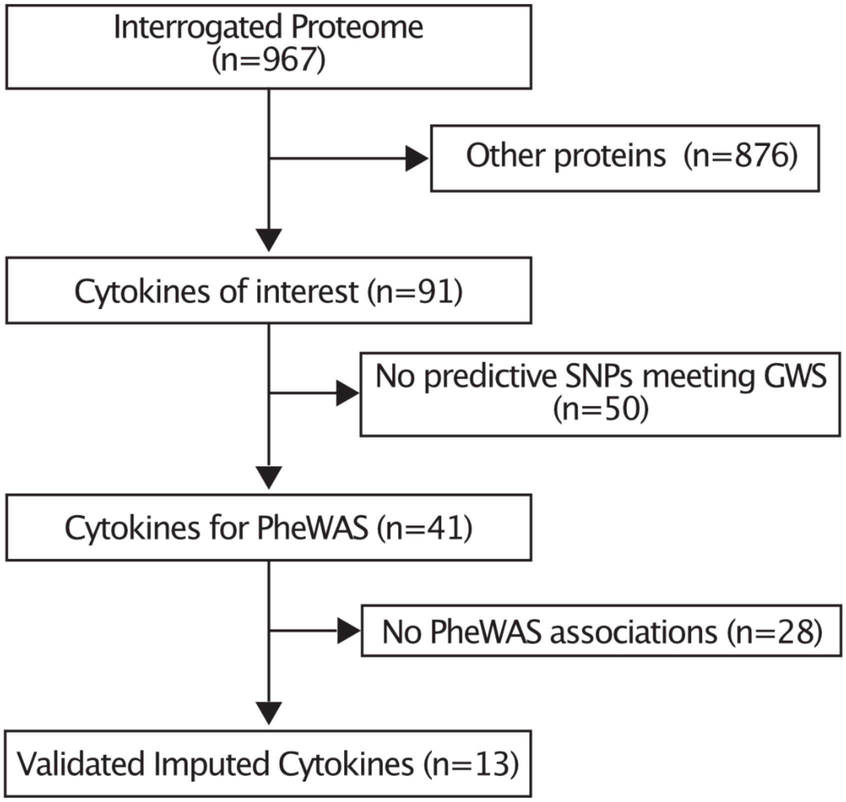


Summary of imputed cytokine validation to use in downstream analyses. Of the available proteome with association summary statistics with single nucleotide polymorphism (SNP), 91 were interleukins, interleukin related circulating proteins, tumor necrosis factors, or tumor necrosis factors related circulating proteins, interferons, interferon related circulating proteins, or C-reactive protein. Of those cytokines of interest, only 41 had SNPs meeting genome wide significance (GWS). Of each of the imputed cytokine level for those 41, Phenome-Wide association studies (PheWAS) were performed using BioVU, the Vanderbilt biobank linked to the SD that contains DNA samples and genetic data derived from clinical blood samples that would otherwise be discarded. Of the imputed cytokines tested, 13 had at least one statistically significant association on PheWAS and were kept for downstream analyses.

### Fig. S3: Feature prevalence at AF diagnosis time by cluster

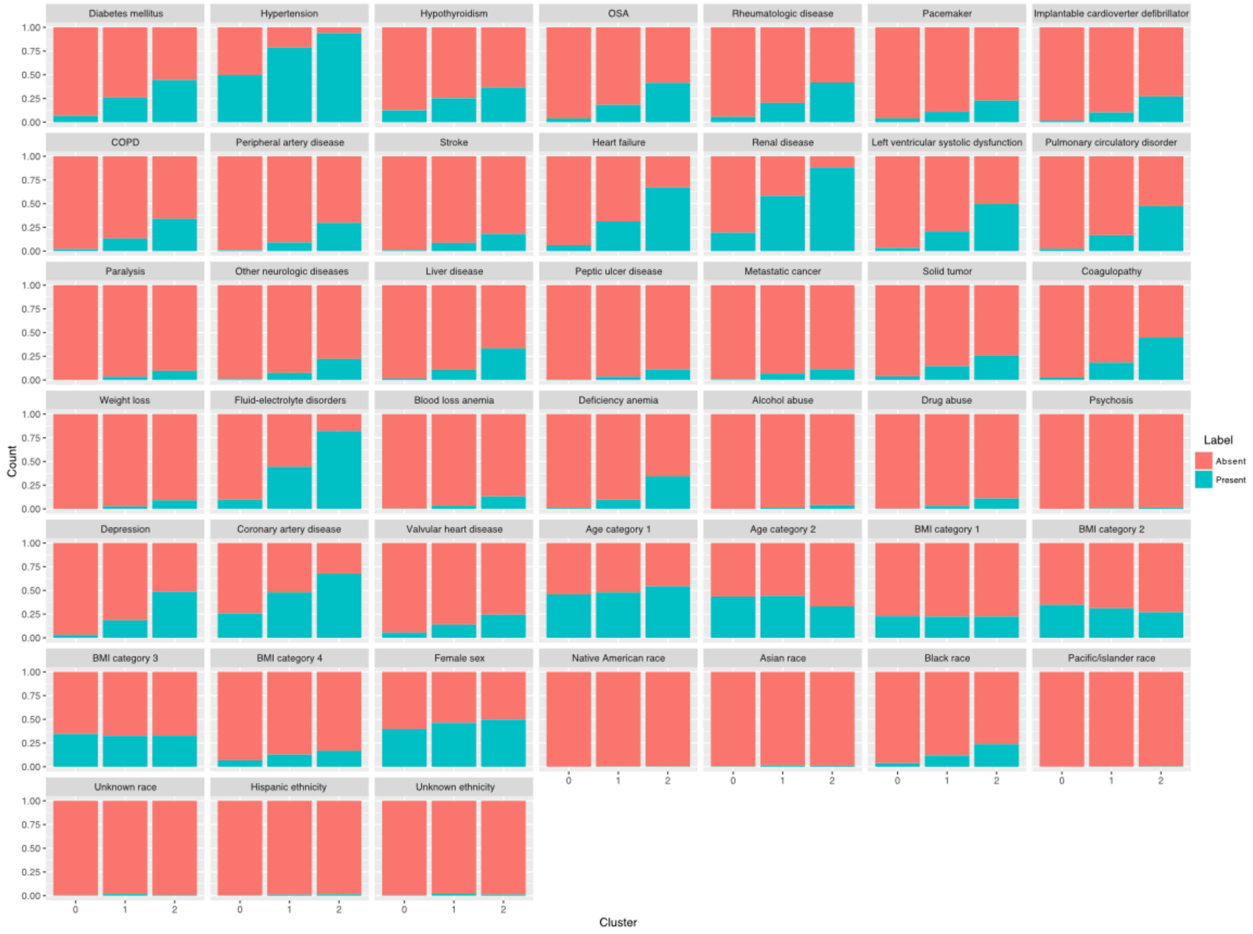


Proportion of additional comorbidities by cluster: Cluster assignment: 0=”Low”, 1=”Mid”, 2=”High”. Abbreviations: OSA: Obstructive sleep apnea, COPD: Chronic obstructive pulmonary disease, BMI: Body mass index.

**Fig. S4: Clinical outcomes during follow up by cluster**

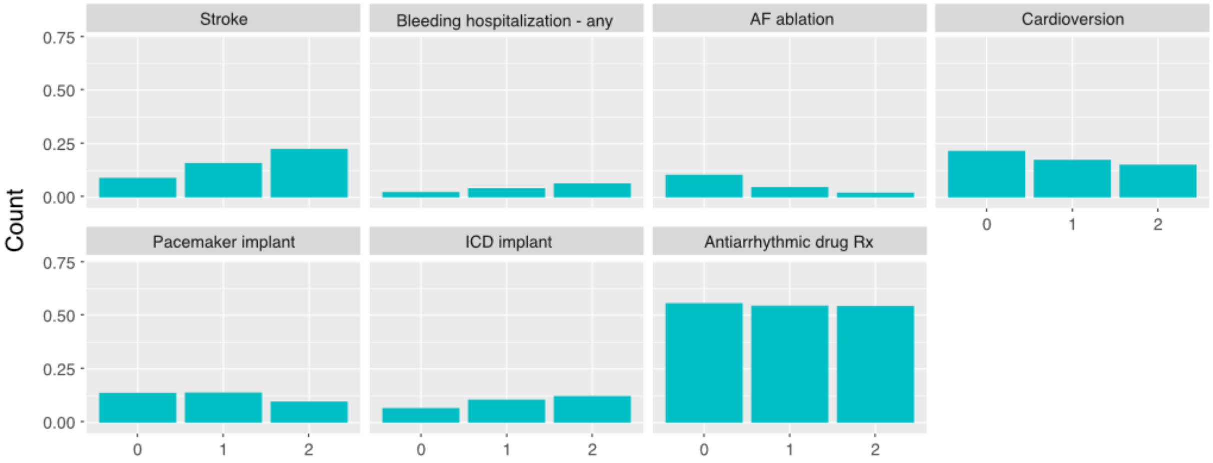


Proportion of outcomes during follow up by cluster: Cluster assignment: 0=”Low”, 1=”Mid”, 2=”High”. Abbreviations: AF: Atrial fibrillation, ICD: Implantable cardioverter defibrillator, Rx: Therapy.
